# Robust Immunohistochemical Detection of α-Synuclein, Tau, and β-Amyloid in Human Brain Tissue Archived for up to 78 Years

**DOI:** 10.64898/2026.02.26.26345861

**Authors:** Mie Kristine Just, Kristina Bang Christensen, Martin Wirenfeldt, Torben Steiniche, Laura Parkkinen, Liisa Myllykangas, Per Borghammer

## Abstract

**Objective:** Brain branks preserve extensive material relevant to neurodegenerative disease research. As these collections age, tissue becomes archival, raising the question of whether long-term fixed and stored human brain tissue remains suitable for contemporary immunohistochemical analyses.

**Materials and Methods:** Forty-one autopsy brains collected between 1946 to 1980 were examined. For each case, midbrain and hippocampus were available both as original paraffin-embedded blocks and as tissue stored long term in fixative. New paraffin blocks were prepared from the long-term fixated tissue. Sections from original and newly prepared blocks were immunohistochemically stained for α-synuclein, hyperphosphorylated tau and amyloid-β. Immunoreactivity was assessed using semi-quantitative scoring.

**Results:** Original blocks consistently showed good staining intensity and morphological preservation for each protein pathology. Newly prepared blocks showed slightly lower semi-quantitative scores for Lewy-related pathology, without statistically significant differences, except for astrocytic α-synuclein in the substantia nigra in cases from the 1960s. Tau pathology displayed modestly reduced labelling, particularly of the neuropil threads and neurofibrillary tangles, most evident in cases from the 1950s. Amyloid-β-positive senile plaques showed similar or slightly higher scores in newly prepared blocks, with no significant differences across regions.

**Conclusion:** Human brain tissue preserved as paraffin-embedded blocks or stored in fixative for up to 78 years remains suitable for immunohistochemical analyses. Adequate-to-good detection of aggregated of α-synuclein, hyperphosphorylated tau and amyloid-β is achievable, indicating preserved pathological hallmarks of Lewy Body Disease and Alzheimer’s Disease in archival tissue.

## Introduction

Alzheimer’s Disease (AD) and Lewy Body Disease (LBD), including Parkinson’s Disease (PD), are the most prevalent neurodegenerative diseases, affecting millions of people worldwide. Neuropathological assessment is primarily based on histological and immunohistochemical (IHC) stains of paraffin-embedded brain tissue. In particular, IHC staining of paraffin-embedded tissue is an invaluable technique used for both diagnostic and research purposes. The pathology of LBD and AD is characterised by dense protein aggregates including intracellular accumulation of α-synuclein (αSyn) into Lewy bodies (LBs) and Lewy neurites (LNs); hyperphosphorylated tau assembling into neurofibrillary tangles (NFTs) and neuropil threads (NTs), and the extracellular accumulation of amyloid-β (Aβ) into senile plaques (SPs).

Staging systems of AD and LBD are based on neuropathological assessment of brain bank material. Systematic brain banking began in the 1940s-1950s [1]. Many early brain bank collections originated from routine hospital autopsies, whereas modern collections often come from donors enrolled in specific brain donation programs. In the 1950s, autopsy rates began to decline steadily, decreasing from 41.1% in 1964 to less than 5% in 1995 [2]. The decline in autopsy rates, and consequently reduced input to brain banks, highlights the importance of assessing and re-assessing archived tissue already available.

In addition to storing formalin-fixed paraffin-embedded (FFPE) blocks, many brain banks and departments of anatomy and pathology also preserve tissue in formalin or similar fixatives. Formalin fixation involves rapid penetration of formaldehyde into the tissue, formation of covalent bonds and cross-linking, i.e. the formation of methylene bridges, with complete fixation typically obtained within 24-48 hours [3-5]. However, fixation is a continuous process, and prolonged fixation may result in reduced antigenicity and IHC efficacy due to structural alterations of epitopes [6] and decreased accessibility of antigens. Furthermore, over longer periods, formalin may oxidize into formic acid (FA), further diminishing antigenicity and compromising IHC results [7].

Common methods for unmasking antigens i.e. antigen retrieval include enzymatic digestion (e.g. proteinase K) or heating in citric acid or FA pretreatment. Some studies have shown that prolonged fixation of brain tissue can compromise IHC staining for αSyn, Aβ and tau, depending on the antibodies used [8-10]. One study investigated prolonged fixation for up to 14 years, and reported reduced antigenicity for αSyn, Aβ, tau, ubiquitin and p62. However, with appropriate combination of antibody and antigen retrieval, antigenicity could be restored for some targets [11].

Antigen preservation and recovery in archival FFPE tissue blocks from 1960 to 2010 have also been investigated. These studies found that most antigens remain well-preserved over several decades. Cytoplasmic antigens were maintained for 60 years or more, while nuclear and membranous antigens were more prone to decay. The study suggested that deeper sectioning and modified pretreatment can optimize antigenicity in old archival FFPE blocks [12].

The Danish Brain Collection contains nearly 10,000 brains from patients admitted to psychiatric hospitals in Denmark, collected between 1945 and 1982 under the Institute of Brain Pathology, Psychiatric Hospital Risskov, Denmark. Approximately 5,500 patients had a diagnosis of dementia, and 1,500 had a diagnosis of schizophrenia. Other types of disorders include manic-depressive disorder, depression, PD, brain tumours, stroke, and more.

The aim of the present study was to evaluate whether very old archival brain material, i.e. both original paraffin-embedded blocks as well as newly sampled tissue with fixation times up to 78 years, remain viable for IHC detection of protein aggregates associated with LBD and AD. The extent of immunoreactive structures were assessed using 41 cases positive for LBD and/or AD protein pathologies, sampled across the decades covered by the Danish Brain Collection.

## Materials and Methods

### Tissue samples

This study was conducted on brain tissue samples obtained from 41 postmortem brains from which the midbrain and the hippocampus were available both as an original paraffin-embedded blocks and in fixative for each case. All included brains displayed Lewy pathology and/or AD pathology. We examined brains that were collected between 1946 and 1980 as part of the brain banking period of the Danish Brain Collection (University of Southern Denmark, Odense). Ethical approval (1-10-72-200-20) was obtained from the Scientific Ethics Committees for the Central Denmark Region.

A minimum of 10 cases were selected from the last half of each decade, i.e. from 1945-1949, 1955-1959, 1965-1969, and 1975-1980. Two cases from the 1940s group were excluded; one due to unusable fixated tissue and one because sections from the original block could not adhere to the slides.

The original blocks had been processed following standard procedures at the time of autopsy and neuropathological assessment and were stored in cardboard boxes at room temperature. To ensure compatibility with our microtome, these original samples were re-embedded in fresh paraffin. The re-embedding procedure involved melting the paraffin of the original block on a warm plate, replacing it with new paraffin in either regular or “mega” moulds depending on the tissue size, and finally mounting new cassettes compatible with the microtome.

Available information on fixative, concentrations and fixation duration was variable. The type of fixative was available in 24 of 43 cases, i.e. 23 brains were formalin-fixed, and one brain was fixed in Lilli’s acetate-formol. For the remaining 19 cases, the fixation type was unknown, though most were presumably fixed in formalin. The duration of fixation of the original blocks is unknown, but standard procedures of the time were followed. Long-fixated brains were stored in fixative in plastic containers at room temperature.

New blocks were sampled from the long-fixated brains, which had previously been cut into approximately 1 cm slices during the original neuropathological evaluation. The new blocks were sampled as close as possible to the original sampling site. Prolonged fixation times of the new blocks were calculated in years from the time of autopsy or death to the year of the new sampling. All tissue samples for this study were sampled between 2020 and 2024. Fixation times of the new blocks and storage times of the original blocks are summarized in **Table 1**.

**Table 1.** Demographics and Tissue Storage Times.

| Decade | Case no. | Age range at death | Sex | Clinical Diagnosis | Postmortem Delay, hours | Fixative | Storage Time of Original Blocks, years | Fixation Time of New Blocks, years |
| --- | --- | --- | --- | --- | --- | --- | --- | --- |
| 1940s | 1 | 56-60 | F | PD | 26 | Formalin | 78 | 76 |
|  | 2 | 71-75 | M | Dementia + PD | N/A | Formalin | 78 | 78 |
|  | 3 | 61-65 | M | Dementia | N/A | Formalin | 77 | 77 |
|  | 4 | 71-75 | F | Dementia | 19 | Formalin | 77 | 75 |
|  | 5 | 76-80 | M | Dementia | 6 | Formalin | 77 | 75 |
|  | 6 | 86-90 | F | Dementia | 24 | Formalin | 76 | 76 |
|  | 7 | 81-85 | M | Dementia | 18 | Formalin | 75 | 75 |
|  | 8 | 76-80 | F | Dementia | 22 | Formalin | 75 | 74 |
|  | 9 | 76-80 | F | Dementia | 8 | Formalin | 74 | N/A |
|  | 10 | 61-65 | M | Dementia + PD | 10 | Formalin | 78 | N/A |
| 1950s | 11 | 66-70 | M | Dementia | 39* | N/A | 69 | 69 |
|  | 12 | 71-75 | F | PD | 25 | Formalin | 69 | 67 |
|  | 13 | 86-90 | M | Dementia + parkinsonism | 7 | Formalin | 69 | 69 |
|  | 14 | 66-70 | F | Dementia + PD | 52 | Formalin | 66 | 66 |
|  | 15 | 86-90 | M | Dementia | 28 | Formalin | 66 | 66 |
|  | 16 | 71-75 | M | Dementia | 10 | N/A | 66 | 66 |
|  | 17 | 66-70 | M | Dementia | 6 | N/A | 66 | 64 |
|  | 18 | 76-80 | M | Dementia + parkinsonism | 14 | Formalin | 66 | 66 |
|  | 19 | 71-75 | M | PD | N/A | N/A | 66 | 66 |
|  | 20 | 71-75 | F | Dementia + parkinsonism | N/A | N/A | 65 | 65 |
|  | 21 | 76-80 | M | Dementia | 113 | Formalin | 65 | 65 |
| 1960s | 22 | 71-75 | M | Dementia + parkinsonism | N/A | N/A | 59 | 59 |
|  | 23 | 66-70 | M | PD | 24* | N/A | 57 | 55 |
|  | 24 | 66-70 | M | Dementia | 7 | N/A | 57 | 57 |
|  | 25 | 46-50 | M | Dementia | 48* | N/A | 57 | 57 |
|  | 26 | 76-80 | F | Dementia | 13 | Formalin | 56 | 56 |
|  | 27 | 66-70 | F | Dementia + parkinsonism | 24* | N/A | 56 | 54 |
|  | 28 | 61-65 | F | Dementia + PD | 16 | Formalin | 56 | 56 |
|  | 29 | 86-90 | F | Dementia | 18 | Formalin | 56 | 54 |
|  | 30 | 71-75 | M | Dementia | N/A | N/A | 55 | 53 |
|  | 31 | 71-75 | F | Dementia + PD | 24 | Formalin | 55 | 55 |
|  | 32 | 81-85 | F | Dementia + PD | N/A | Formalin | 55 | 55 |
|  | 33 | 76-80 | M | PD | 48* | N/A | 55 | 55 |
| 1970s | 34 | 86-90 | M | Dementia | 24* | Formalin | 49 | 47 |
|  | 35 | 71-75 | M | Dementia | 20* | N/A | 49 | 49 |
|  | 36 | 76-80 | F | Dementia | N/A | N/A | 48 | 46 |
|  | 37 | 71-75 | F | Dementia + parkinsonism | N/A | N/A | 46 | 42 |
|  | 38 | 71-75 | F | Dementia | N/A | N/A | 45 | 41 |
|  | 39 | 71-75 | F | Dementia | 22* | Formalin | 45 | 41 |
|  | 40 | 76-80 | F | Dementia + PD | N/A | N/A | 45 | 41 |
|  | 41 | 76-80 | M | Dementia + PD | N/A | N/A | 44 | 40 |
|  | 42 | 76-80 | F | Manic-depressive psychosis | 48* | Lilli's Acetate-formol | 44 | 42 |
|  | 43 | 71-75 | M | Dementia + PD | N/A | N/A | 44 | 40 |
|  |  | Avg. age | % M |  | Avg. PMD |  | Avg. storage time | Avg. fixation time |
| 1940s | n = 8 | 74.4 | 50 |  | 16.6 |  | 76.6 | 75.8 |
| 1950s | n = 11 | 75.5 | 80 |  | 32.7 |  | 66.6 | 66.3 |
| 1960s | n = 12 | 71.6 | 60 |  | 24.7 |  | 56.2 | 55.5 |
| 1970s | n = 10 | 76.4 | 40 |  | 28.5 |  | 45.9 | 42.9 |

\* PMD is an approximation as only the date of autopsy is known. Abbreviations: AD, Alzheimer's Disease; avg., average; N/A, not available; PD, Parkinson's Disease; PMD, postmortem delay.

Based on the minimum recommended brain regions to be sampled by Montine and colleagues [13] and the regions originally sampled, the following areas were evaluated: the midbrain including the substantia nigra (SN) and periaqueductal grey (PAG), and the hippocampal block, including CA1 and CA2 of the hippocampus and the entorhinal cortex (Ent Cx) and the temporo-occipital cortex (TO Cx). Details on which regions were analysed for specific types of pathology are provided in **Table 2**.

**Table 2.** Regions Analysed for Each Type of Pathology.

| Pathology | Midbrain | Hippocampus | Cortex |
| --- | --- | --- | --- |
| Amyloid plaques | PAG | CA1 and CA2 | Ent Cx + TO Cx |
| Cerebral Amyloid Angiopathy |  |  | Ent Cx + TO Cx |
| NFTs |  | CA1 and CA2 | Ent Cx + TO Cx |
| NTs |  | CA1 and CA2 | Ent Cx + TO Cx |
| LBs (LB-like inclusions*) | SN | CA2 | Ent Cx + TO Cx |
| LNs | SN | CA2 | Ent Cx + TO Cx |
| Astrocytic αSyn | SN | CA2 | Ent Cx + TO Cx |

\*Intracellular, intraneuritic and extracellular inclusions

### Immunohistochemistry

Immunohistochemical staining was performed with an automated Ventana Benchmark Ultra immunostainer (Ventana Medical Systems, Arizona, USA) to standardise the technique. Tissue sections of 4 µm were deparaffinized and subjected to antigen retrieval using Envision Flex Target Retrieval Solution (DAKO) in a pressure cooker (decloaking chamber™ NxGen Manual, Biocare Medical, California, USA) at 95ºC, followed by incubation in 98% formic acid (VWR, Avantor, Pennsylvania, USA). Details on antigen retrieval are provided in **Table 3**. The remaining immunohistochemical staining steps were performed on the Ventana Benchmark Ultra (Ventana Medical Systems, Arizona, USA). Based on previous IHC studies on archival brain tissue, the non-commercial 5G4 antibody was chosen for detecting αSyn as it had shown promising IHC results on long-fixed brain tissue. Sections were stained for αSyn (5G4 #847-0102004001, 1:5000, AJ Roboscreen GmbH Leipzig, Germany), Aβ (4G8 #9220-02, 1:14000, BioLegend, California, USA) and hyperphosphorylated tau (AT8 #90206, 1:1000, Fujirebio, Pennsylvania, USA). Visualization was achieved using Optiview DAB (Roche Diagnostics, Ventana, Arizona, USA), and haematoxylin (Roche Diagnostics, Ventana, Arizona, USA) was used for counterstaining.

**Table 3.** Immunohistochemistry: Antibodies and Antigen Retrieval.

| Antibody | Clone | Vendor | Dilution | Target Retrieval | Formic Acid |
| --- | --- | --- | --- | --- | --- |
| αSyn | 5G4 | Roboscreen | 1:5000 | 30 min, 95°C, pH = 6 | 30 min |
| HP-Tau | AT8 | Fujirebio | 1:1000 | 20 min, 95 °C, pH = 9 | 5 min |
| Aβ | 4G8 | BioLegend | 1:14,000 | 20 min, 95 °C, pH = 9 | 5 min |

αSyn, alpha-synuclein; Aβ , amyloid-beta; HP-Tau, hyperphosphorylated tau

### Digital assessment

Whole slide scans of sections from the midbrain, hippocampal blocks including Ent Cx and TO Cx were captured on a digital slide scanner (S60, Hamamatsu, Hamamatsu City, Japan). Circular regions of interest (ROIs) of 2 mm diameter were assessed digitally at x20 magnification. The ROIs were placed at the hotspot of pathology within the assessed region.

### Assessment of pathology

All samples were analysed by the same evaluator. The extent of αSyn, tau and Aβ pathologies were semi-quantitatively assessed at a magnification of x20 as previously described [11]. In short, intracellular inclusions (LBs and NFTs) and extracellular aggregates (SPs) were scored: 0, no lesions; 1, 1 to 5 lesions; 2, 6 to 20 lesions; 3, more than 20 lesions. The neuritic densities of LNs and NTs and astrocytic density of αSyn were assessed on a semiquantitative scale of 0-3: 0, absent; 1, occasional; 2, moderate; 3, severe. Briefly, occasionally present meant LNs/NTs/astrocytic αSyn had to be sought out. Densities were scored moderate, when they were readily seen. Lastly, densities were rated severe when numerous LNs/NTs/αSyn-positive astrocytes were present. The presence or absence of cerebral amyloid angiopathy (CAA) was noted.

Staining quality was estimated on a 3-step scale (good, acceptable, or poor). Briefly, the staining quality was assessed as good when the pathological structures were clearly labelled with no or very faint background stain. The staining was acceptable when structures were partially labelled but still detectable or were well-labelled but showed some background staining. The staining was assessed as poor if labelling was faint or the region showed excessive background staining. **Table 4** summarises the assessment of staining quality.

**Table 4.**
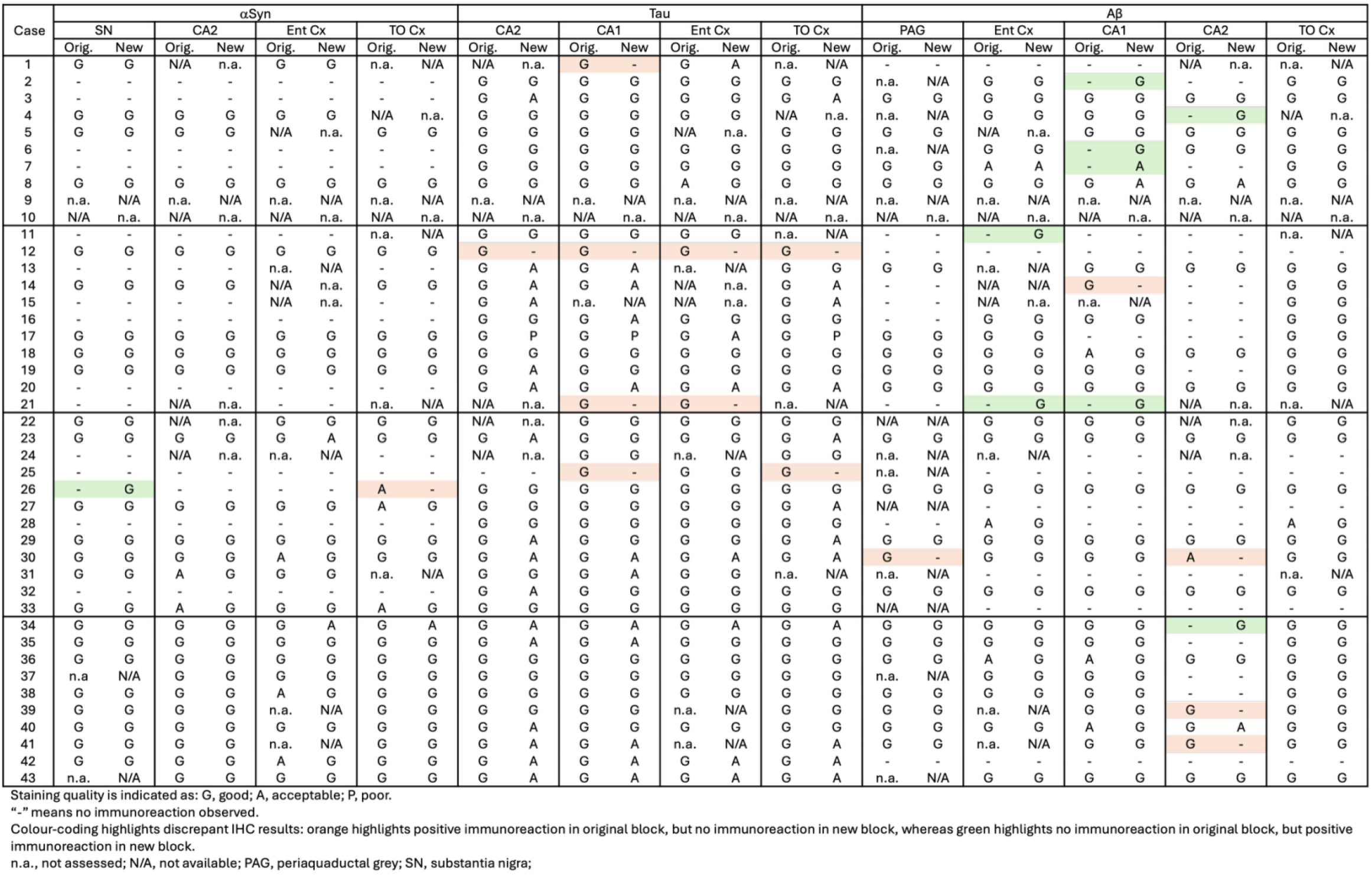
Staining Quality of Immunohistochemical Stains.

### Statistical Analysis

The statistical analyses were performed using GraphPad Prism version 10.6.1 for macOS, GraphPad Software, Boston, Massachusetts USA. The comparison of semiquantitative assessments of the two types of preserved tissue was estimated applying the nonparametric Wilcoxon rank-sum test. P values are shown in tables by asterisks: *, p < 0.05, **, p < 0.01.

## Results

A total of 162 paraffin-embedded blocks were included, 394 IHC stains were performed and 944 regions were semi-quantitatively assessed. In general, good staining quality was achieved across all decades, ranging from 1946 to 1980, and across both types of tissue preservation, i.e. original blocks stored in cardboard boxes for up to 78 years and newly sampled blocks with fixation times up to 78 years. In the comparison below, the original blocks were regarded as the gold standard, and the antigenicity of new blocks was evaluated relative to that of original blocks. Representative images of the staining quality of αSyn, tau, and Aβ pathology across the decades are provided in **Fig. 1-5**.

**Figure 1.**
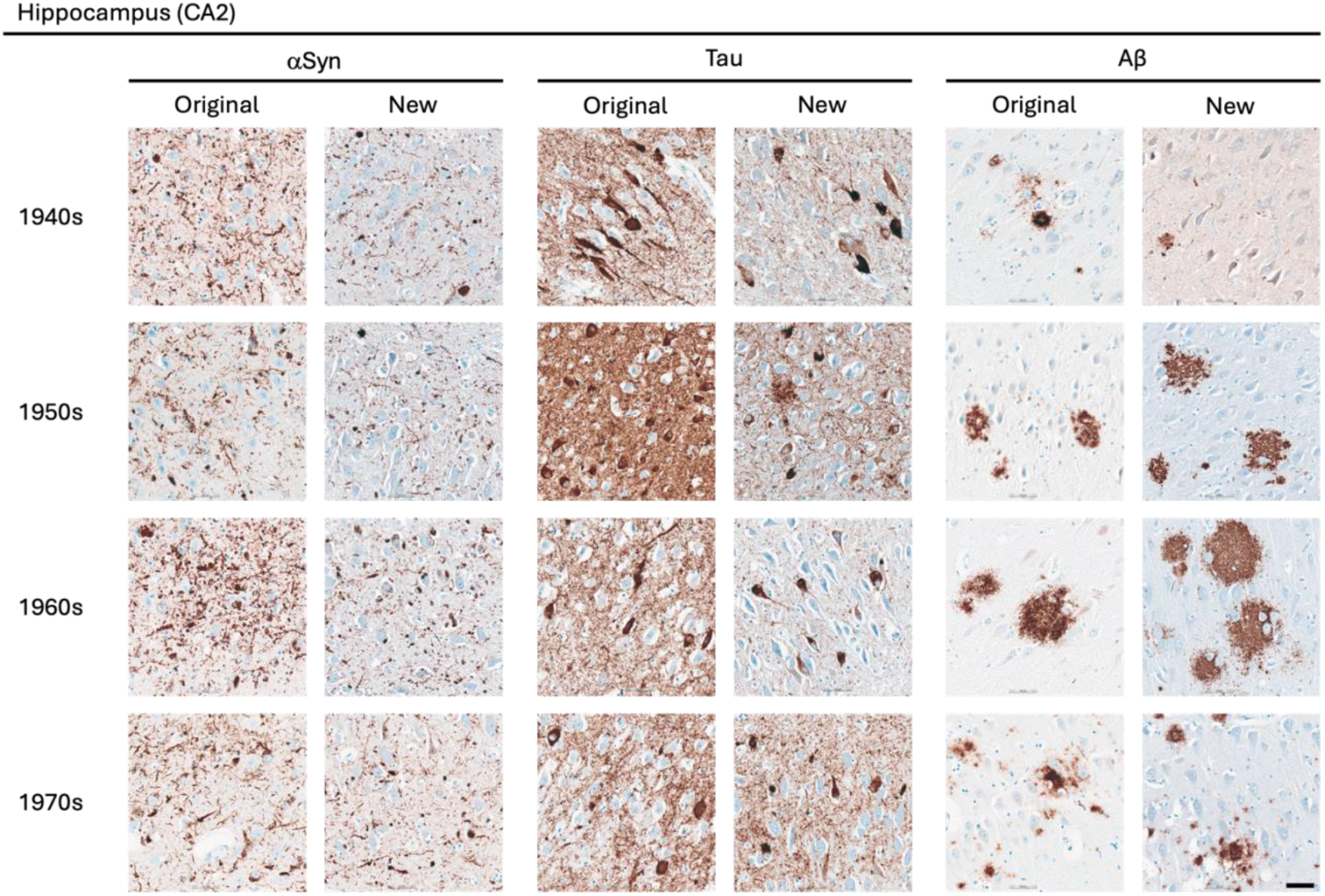
Representative images of immunohistochemical stains of CA2 of the hippocampus for originally paraffin-embedded blocks (Original) and for newly paraffin-embedded blocks upon prolonged fixation (New). **αSyn** Photomicrographs comparing 5G4 IHC staining seen in same case per decade. **Tau** Photomicrographs comparing AT8 IHC staining seen in same case per decade. Note that intense staining represents specific tau staining. **Aβ** Photomicrographs comparing 4G8 IHC staining seen in same case per decade. Scale bar = 50 μm.

**Figure 2.**
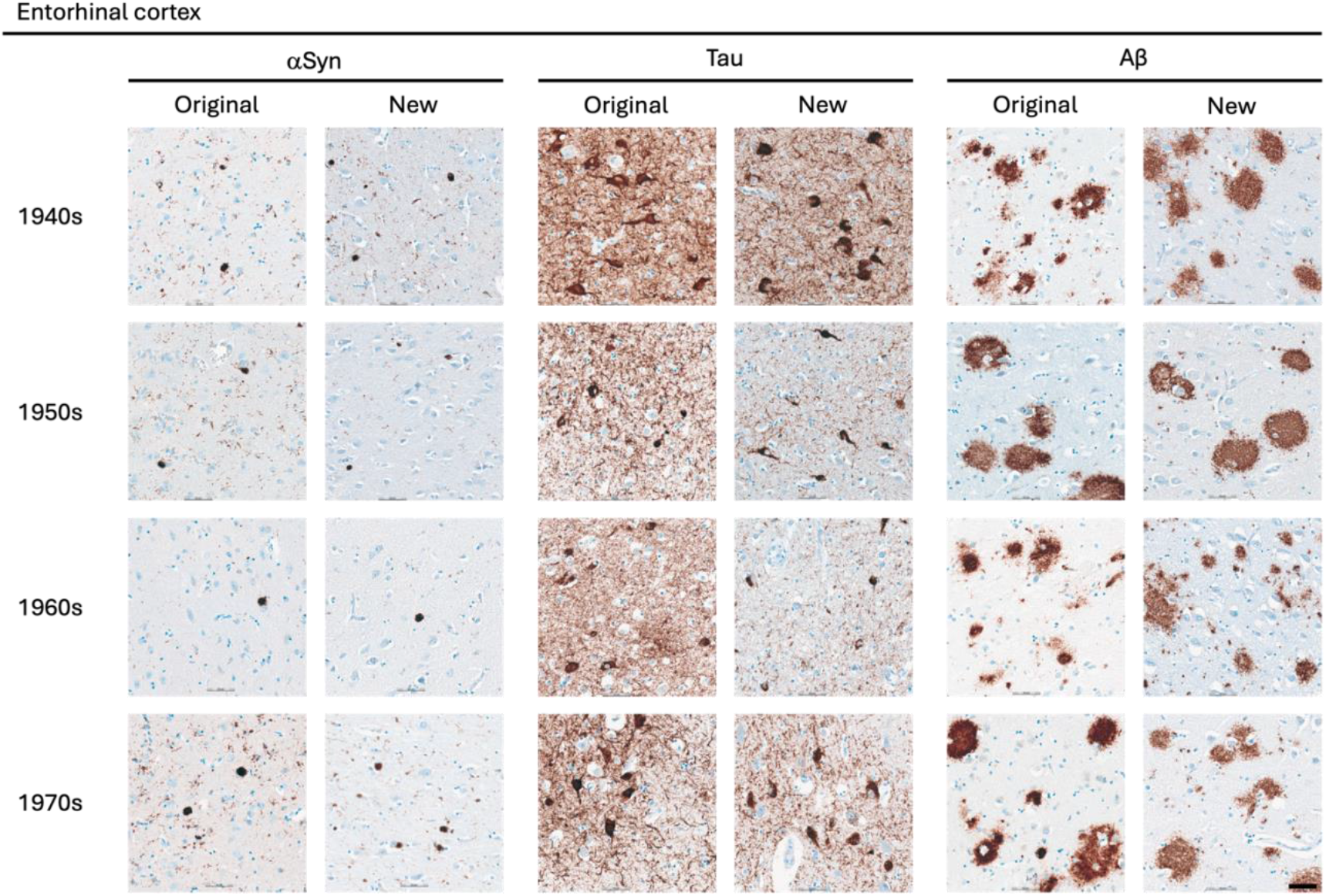
Representative images of immunohistochemical stains of entorhinal cortex for originally paraffin-embedded blocks (Original) and for newly paraffin-embedded blocks upon prolonged fixation (New). **αSyn** Photomicrographs comparing 5G4 IHC staining seen in same case per decade. **Tau** Photomicrographs comparing AT8 IHC staining seen in same case per decade. **Aβ** Photomicrographs comparing 4G8 IHC staining seen in same case per decade. Scale bar = 50 μm.

**Figure 3.**
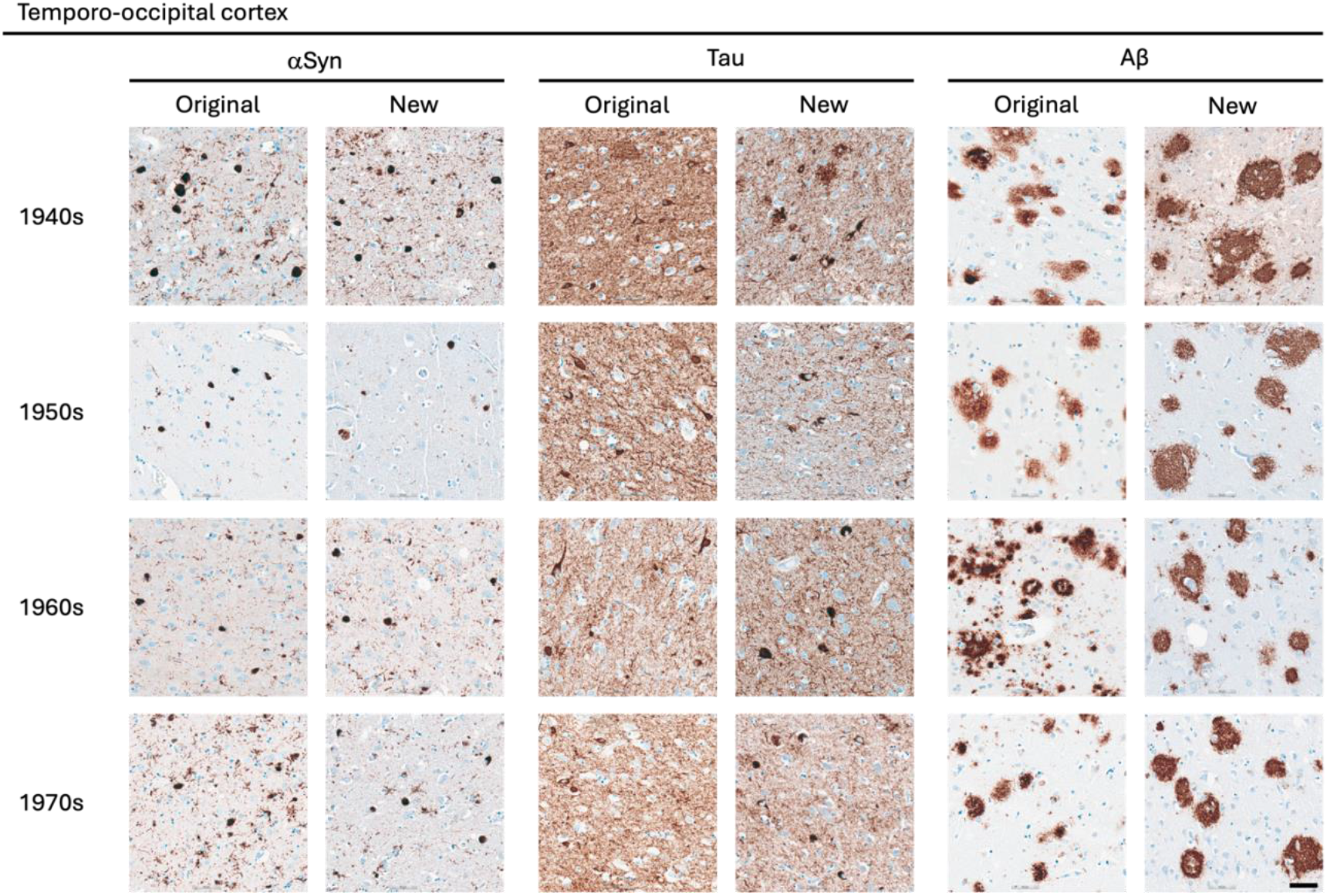
Representative images of immunohistochemical stains of temporo-occipital cortex for originally paraffin-embedded blocks (Original) and for newly paraffin-embedded blocks upon prolonged fixation (New). **αSyn** Photomicrographs comparing 5G4 IHC staining seen in same case per decade. **Tau** Photomicrographs comparing AT8 IHC staining seen in same case per decade. **Aβ**Photomicrographs comparing 4G8 IHC staining seen in same case per decade. Scale bar = 50 μm.

**Figure 4.**
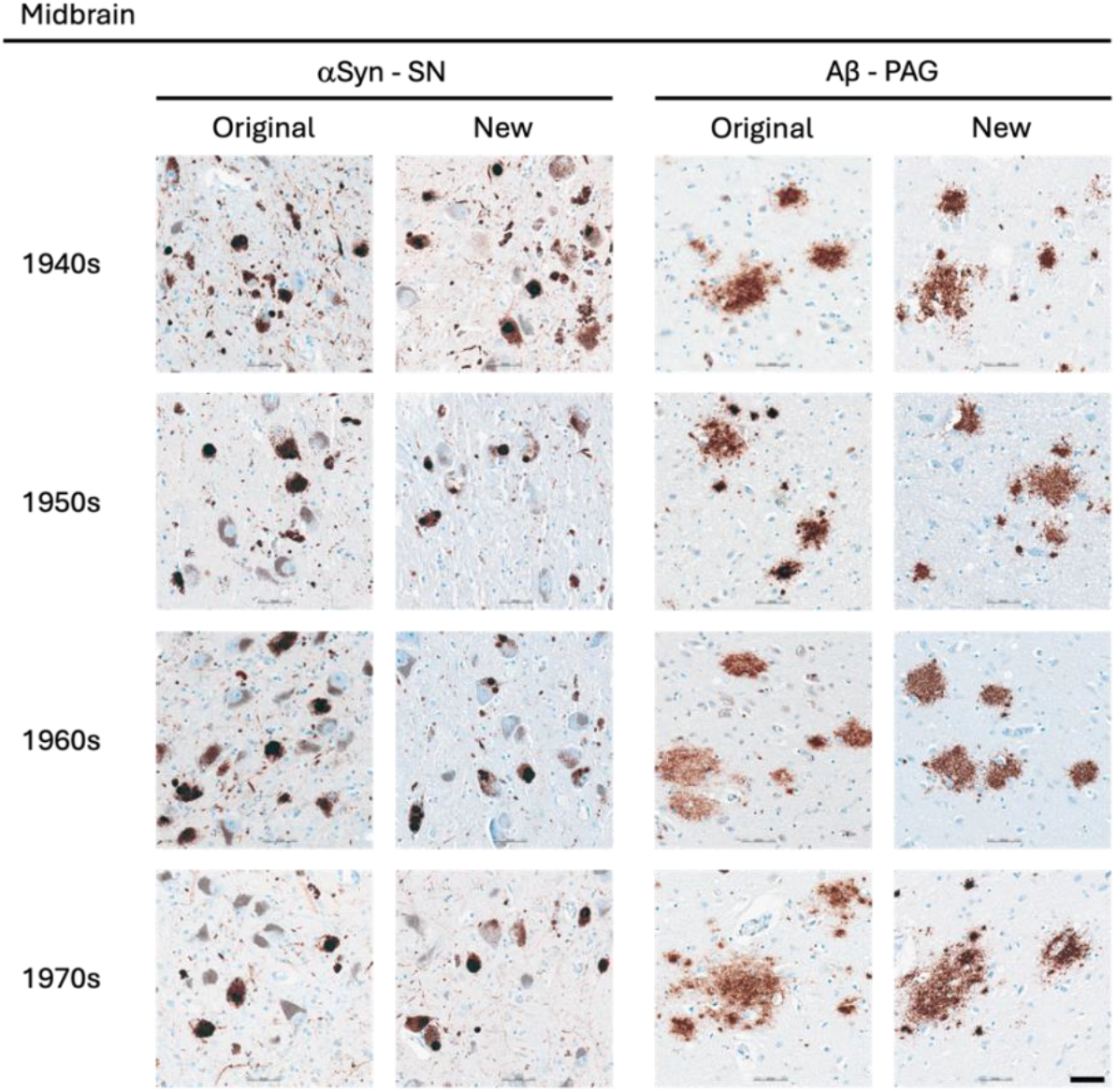
Representative images of immunohistochemical stains of midbrain for originally paraffin-embedded blocks (Original) and for newly paraffin-embedded blocks upon prolonged fixation (New). **αSyn** Lewy pathology labelled by 5G4 IHC in the SN across the four decades. **Aβ** Senile plaques detected with 4G8 IHC in original and new blocks across the decades. Scale bar =50 μm. PAG, periaqueductal grey; SN, substantia nigra

**Figure 5.**
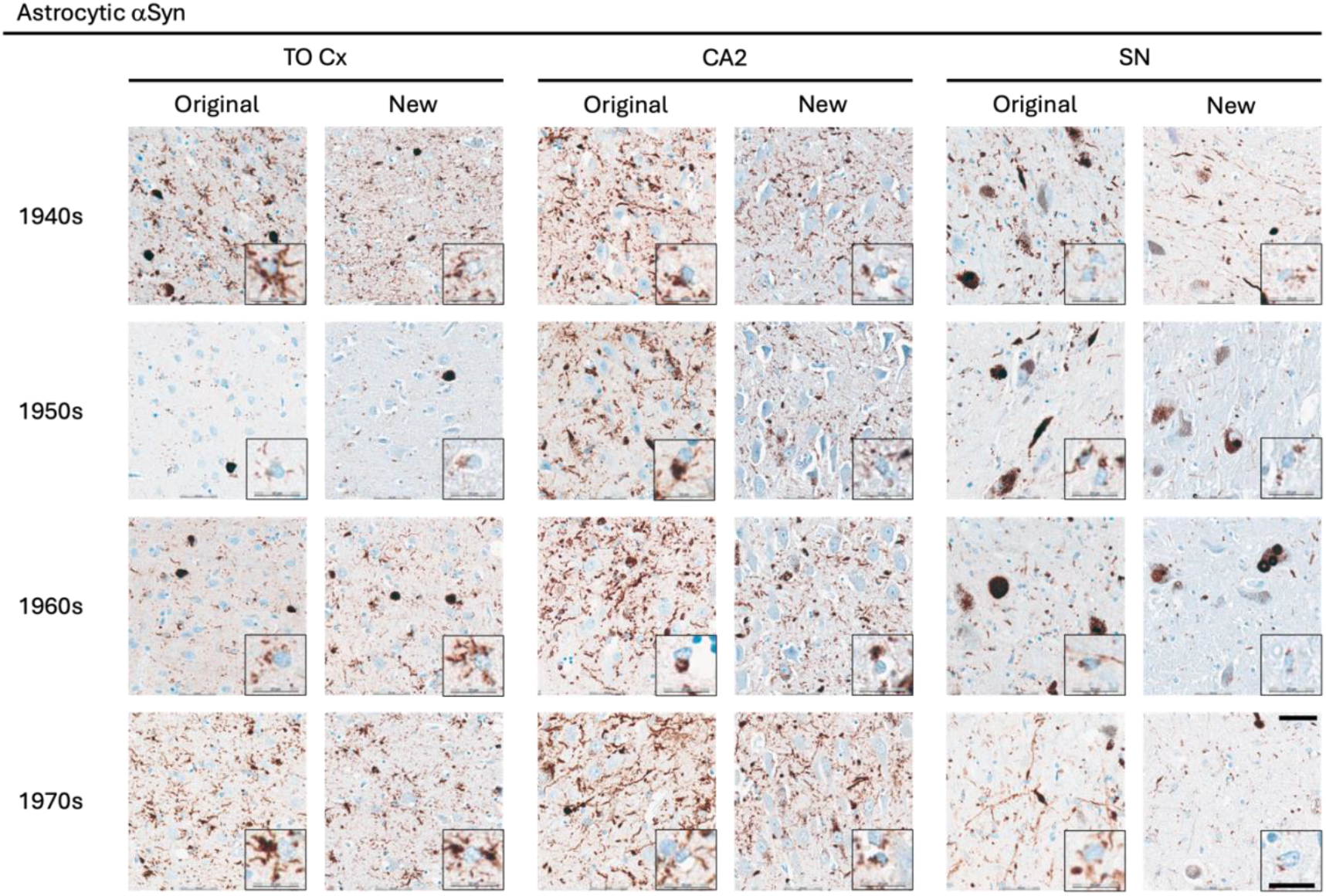
Representative images of immunohistochemical stains of astrocytic **α**Syn in temporo-occipital cortex, CA2 of the hippocampus and substantia nigra for originally paraffin-embedded blocks (Original) and for newly paraffin-embedded blocks upon prolonged fixation (New). Inserts show magnification of astrocytes containing **α**Syn labelled with the 5G4 antibody. Scale bars = 50 μm and 20 μm for inserts. SN, substantia nigra; TO Cx, temporo-occipital cortex.

### αSyn

The 5G4 antibody labelled both LBs and LNs with good staining quality and intensity in the original blocks. Comparable results were obtained in the new blocks with long fixation times (**Fig. 1-4**). Numerically, the new blocks showed slightly lower semi-quantitative scores than the original blocks, but no statistically significant differences were observed in any region **Table 5**. Astrocytic αSyn aggregates were generally adequately labelled (**Fig. 5**) and these also showed slightly lower numerical scores on average across most regions, but a statistical significant difference was seen only in the substantia nigra of the 1960s group (p < 0.05) (**Table 5**).

**Table 5.** Semiquantitative Assessment of αSyn Pathology.

| αSyn |  | Lewy Bodies |  |  | Lewy Neurites |  |  | Astrocytic αSyn |  |  |
| --- | --- | --- | --- | --- | --- | --- | --- | --- | --- | --- |
|  | Sample size | Semiquantitative assessment |  |  | Semiquantitative assessment |  |  | Semiquantitative assessment |  |  |
|  |  | Original Mean ± SEM | New Mean ± SEM | Wilcoxon test | Original Mean ± SEM | New Mean ± SEM | Wilcoxon test | Original Mean ± SEM | New Mean ± SEM | Wilcoxon test |
| **SN** |  |  |  |  |  |  |  |  |  |  |
| 1940s | n = 8 | 1.3 ± 0.5 | 0.9 ± 0.4 | ns | 1.4 ± 0.5 | 1.0 ± 0.4 | ns | 1.1 ± 0.5 | 0.8 ± 0.3 | ns |
| 1950s | n = 11 | 0.9 ± 0.4 | 0.8 ± 0.3 | ns | 0.9 ± 0.3 | 0.7 ± 0.3 | ns | 0.9 ± 0.3 | 0.6 ± 0.2 | ns |
| 1960s | n = 12 | 1.5 ± 0.4 | 1.0 ± 0.3 | ns | 1.4 ± 0.4 | 1.0 ± 0.3 | ns | 1.7 ± 0.4 | 1.1 ± 0.3 | * |
| 1970s | n = 8 | 2.4 ± 0.2 | 1.9 ± 0.1 | ns | 2.1 ± 0.1 | 1.9 ± 0.1 | ns | 2.1 ± 0.1 | 2.3 ± 0.2 | ns |
| **CA2** |  |  |  |  |  |  |  |  |  |  |
| 1940s | n = 7 | 0.7 ± 0.4 | 0.6 ± 0.3 | ns | 1.0 ± 0.5 | 0.9 ± 0.4 | ns | 1.1 ± 0.6 | 1.1 ± 0.6 | ns |
| 1950s | n = 10 | 0.3 ± 0.2 | 0.2 ± 0.1 | ns | 0.7 ± 0.3 | 0.6 ± 0.2 | ns | 0.9 ± 0.4 | 0.8 ± 0.4 | ns |
| 1960s | n = 10 | 0.6 ± 0.2 | 0.3 ± 0.2 | ns | 1.1 ± 0.4 | 0.7 ± 0.2 | ns | 1.3 ± 0.4 | 0.9 ± 0.4 | ns |
| 1970s | n = 10 | 0.8 ± 0.3 | 0.9 ± 0.2 | ns | 2.0 ± 0.3 | 1.7 ± 0.2 | ns | 2.3 ± 0.3 | 2.1 ± 0.4 | ns |
| **Ent Cx** |  |  |  |  |  |  |  |  |  |  |
| 1940s | n = 7 | 0.7 ± 0.4 | 0.9 ± 0.5 | ns | 0.7 ± 0.4 | 0.9 ± 0.5 | ns | 1.0 ± 0.5 | 1.0 ± 0.5 | ns |
| 1950s | n = 8 | 0.4 ± 0.3 | 0.4 ± 0.2 | ns | 0.8 ± 0.3 | 0.5 ± 0.2 | ns | 1.0 ± 0.4 | 0.5 ± 0.2 | ns |
| 1960s | n = 11 | 1.2 ± 0.3 | 0.9 ± 0.3 | ns | 1.2 ± 0.3 | 0.8 ± 0.2 | ns | 1.5 ± 0.4 | 1.2 ± 0.3 | ns |
| 1970s | n = 8 | 1.6 ± 0.2 | 1.5 ± 0.3 | ns | 1.8 ± 0.2 | 1.8 ± 0.3 | ns | 2.5 ± 0.3 | 2.3 ± 0.4 | ns |
| **TO Cx** |  |  |  |  |  |  |  |  |  |  |
| 1940s | n = 6 | 1.0 ± 0.6 | 0.8 ± 0.5 | ns | 1.0 ± 0.6 | 0.8 ± 0.5 | ns | 1.0 ± 0.6 | 1.0 ± 0.6 | ns |
| 1950s | n = 9 | 0.8 ± 0.3 | 0.7 ± 0.3 | ns | 0.9 ± 0.3 | 0.7 ± 0.2 | ns | 1.0 ± 0.3 | 0.9 ± 0.4 | ns |
| 1960s | n = 11 | 1.1 ± 0.4 | 0.9 ± 0.3 | ns | 1.2 ± 0.4 | 0.8 ± 0.3 | ns | 1.3 ± 0.4 | 0.9 ± 0.4 | ns |
| 1970s | n = 10 | 2.3 ± 0.2 | 2.1 ± 0.3 | ns | 2.4 ± 0.2 | 1.9 ± 0.2 | ns | 2.7 ± 0.2 | 2.5 ± 0.2 | ns |

ns, non-significant; \*, $p < 0.05$

### Tau

The AT8 antibody labelled NFTs and NTs with good staining quality and intensity in the original blocks across the decades (**Fig. 1-3**). In the 1940s group, the new blocks showed semi-quantitative scores comparable to the original blocks, with some slightly lower and some slightly higher, but no significant differences were observed in any region. The 1950s group displayed the greatest difference between sections from original and new blocks. Significant differences in semi-quantitative scores were found for NTs across all regions (CA2, p < 0.05; CA1, p < 0.01; Ent Cx, p < 0.05; TO Cx, p < 0.05), whereas for NFTs, significant differences were observed only in the CA2 region of the hippocampus (p < 0.05) and the TO Cx (p < 0.05). In the 1960s group, new blocks showed slightly lower semi-quantitative scores than original blocks, but no significant differences were detected for NFTs in any region. Labelling of NTs also showed slightly lower numerical scores on average, with significant differences detected only in the CA1 of the hippocampus (p < 0.05) and the Ent Cx (p < 0.05). In the 1970s group, new blocks again showed slightly lower semi-quantitative scores on average across most regions, with a significant difference seen only for NTs in the CA1 of the hippocampus (p < 0.05) (**Table** 6).

**Table 6.** Semiquantitative Assessment of Tau Pathology.

| Tau |  | Neurofibrillary Tangles |  |  | Neuropil Threads |  |  |
| --- | --- | --- | --- | --- | --- | --- | --- |
|  | Sample size | Semiquantitative assessment |  |  | Semiquantitative assessment |  |  |
|  |  | Original<br>Mean ± SEM | New<br>Mean ± SEM | Wilcoxon test | Original<br>Mean ± SEM | New<br>Mean ± SEM | Wilcoxon test |
| CA2 |  |  |  |  |  |  |  |
| 1940s | n = 7 | 2.4 ± 0.3 | 1.9 ± 0.4 | ns | 2.9 ± 0.1 | 2.4 ± 0.3 | ns |
| 1950s | n = 10 | 1.9 ± 0.4 | 0.9 ± 0.3 | * | 2.3 ± 0.3 | 1.1 ± 0.2 | * |
| 1960s | n = 10 | 1.3 ± 0.5 | 0.7 ± 0.3 | ns | 1.7 ± 0.4 | 1.3 ± 0.2 | ns |
| 1970s | n = 10 | 2.0 ± 0.3 | 1.3 ± 0.3 | * | 2.4 ± 0.3 | 1.8 ± 0.3 | ns |
| CA1 |  |  |  |  |  |  |  |
| 1940s | n = 8 | 2.4 ± 0.3 | 2.4 ± 0.4 | ns | 2.4 ± 0.3 | 2.3 ± 0.4 | ns |
| 1950s | n = 10 | 1.9 ± 0.3 | 1.2 ± 0.4 | ns | 2.1 ± 0.3 | 1.1 ± 0.3 | ** |
| 1960s | n = 12 | 1.6 ± 0.4 | 1.3 ± 0.4 | ns | 1.8 ± 0.3 | 1.3 ± 0.2 | * |
| 1970s | n = 10 | 2.4 ± 0.2 | 2.0 ± 0.4 | ns | 2.3 ± 0.3 | 1.6 ± 0.2 | * |
| Ent Cx |  |  |  |  |  |  |  |
| 1940s | n = 7 | 2.1 ± 0.5 | 2.1 ± 0.3 | ns | 2.3 ± 0.4 | 2.0 ± 0.3 | ns |
| 1950s | n = 8 | 1.9 ± 0.4 | 1.0 ± 0.4 | ns | 2.1 ± 0.4 | 1.0 ± 0.3 | * |
| 1960s | n = 11 | 1.9 ± 0.3 | 1.4 ± 0.4 | ns | 2.1 ± 0.3 | 1.5 ± 0.2 | * |
| 1970s | n = 8 | 2.3 ± 0.3 | 1.9 ± 0.4 | ns | 2.4 ± 0.3 | 1.9 ± 0.3 | ns |
| TO Cx |  |  |  |  |  |  |  |
| 1940s | n = 6 | 2.3 ± 0.3 | 2.7 ± 0.2 | ns | 2.5 ± 0.3 | 2.8 ± 0.2 | ns |
| 1950s | n = 10 | 2.1 ± 0.3 | 1.2 ± 0.3 | * | 2.1 ± 0.3 | 1.2 ± 0.3 | * |
| 1960s | n = 11 | 1.5 ± 0.4 | 1.2 ± 0.4 | ns | 1.6 ± 0.3 | 1.3 ± 0.2 | ns |
| 1970s | n = 10 | 1.9 ± 0.3 | 1.8 ± 0.3 | ns | 2.2 ± 0.3 | 1.9 ± 0.3 | ns |

ns, non-significant; \*, $p < 0.05$ ; \*\*, $p < 0.01$

### Aβ

The 4G8 antibody reliably detected senile plaques in both types of preserved tissue, with good staining intensity and quality across all decades (**Fig. 1-4**). Comparisons between original and new blocks showed similar or slightly higher numerical scores in the new blocks across the decades and anatomical regions, but no significant differences were detected (**Table 7**). Cerebral amyloid angiopathy (CAA) was observed in only 6 of 41 cases, but in these cases, CAA was present in both cortical regions of the original and new blocks (data not shown).

**Table 7.** Semiquantitative Assessment of Amyloid-β Pathology.

| **Aβ** | Senile Plaques |  |  |  |
| --- | --- | --- | --- | --- |
|  | Semiquantitative assessment |  |  |  |
| | Sample size | Original<br>Mean ± SEM | New<br>Mean ± SEM | Wilcoxon test |
| **Ent Cx** |  |  |  |  |
| 1940s | n = 7 | 2.3 ± 0.5 | 2.4 ± 0.4 | ns |
| 1950s | n = 8 | 1.6 ± 0.5 | 2.3 ± 0.4 | ns |
| 1960s | n = 11 | 1.7 ± 0.5 | 1.8 ± 0.4 | ns |
| 1970s | n = 8 | 2.6 ± 0.4 | 2.6 ± 0.4 | ns |
| **CA1** |  |  |  |  |
| 1940s | n = 8 | 1.3 ± 0.5 | 2.0 ± 0.4 | ns |
| 1950s | n = 10 | 1.4 ± 0.4 | 1.3 ± 0.4 | ns |
| 1960s | n = 12 | 1.0 ± 0.3 | 1.2 ± 0.4 | ns |
| 1970s | n = 10 | 1.9 ± 0.2 | 2.4 ± 0.3 | ns |
| **CA2** |  |  |  |  |
| 1940s | n = 7 | 0.9 ± 0.3 | 0.9 ± 0.3 | ns |
| 1950s | n = 10 | 0.5 ± 0.3 | 0.5 ± 0.3 | ns |
| 1960s | n = 10 | 0.7 ± 0.3 | 0.7 ± 0.3 | ns |
| 1970s | n = 10 | 0.9 ± 0.4 | 0.5 ± 0.2 | ns |
| **TO Cx** |  |  |  |  |
| 1940s | n = 6 | 2.7 ± 0.2 | 2.8 ± 0.2 | ns |
| 1950s | n = 9 | 2.7 ± 0.3 | 2.7 ± 0.3 | ns |
| 1960s | n = 11 | 1.8 ± 0.4 | 1.8 ± 0.4 | ns |
| 1970s | n = 10 | 2.7 ± 0.3 | 2.7 ± 0.3 | ns |
| **PAG** |  |  |  |  |
| 1940s | n = 5 | 1.6 ± 0.5 | 2.0 ± 0.6 | ns |
| 1950s | n = 11 | 1.2 ± 0.4 | 1.2 ± 0.4 | ns |
| 1960s | n = 6 | 1.5 ± 0.5 | 1.7 ± 0.6 | ns |
| 1970s | n = 8 | 1.4 ± 0.3 | 1.8 ± 0.3 | ns |

ns, non-significant

## Discussion

Some very old brain collections featured obsolete tissue handling such as formalin fixation protocols spanning years or decades as was the case in this study. Yet, these old collections represent a unique scientific resource, since the brain donors lived during a different era, for example prior to exposure to modern environmental toxicants and before the advent of modern types of commonly used medication. Here, we assessed αSyn, tau and Aβ pathology in long-term stored paraffin-embedded blocks and in prolonged fixated tissue with preservation times ranging from 45 to 78 years. To our knowledge, this is the first systematic study evaluating such old brain tissue with respect to antigenicity of protein aggregates present in AD and LBD. Our findings demonstrate that aggregates of αSyn, tau, and Aβ can be satisfactorily labelled with immunohistochemical techniques with optimised antigen retrieval methods.

Previous studies have reported challenges in detecting αSyn in long-fixated tissue. Pikkarainen and colleagues found that only clone 42, a commercial antibody against αSyn, detected αSyn-immunoreactive structures in tissue fixated for a long time [11]. Kovacs and colleagues introduced the 5G4 antibody, which labelled more neuritic and intracellular structures compared to clone 42 in archival tissue fixed up to 14 years. All antibodies but 5G4 in that study showed background and/or normal synaptic staining, also known as synaptophysin-like staining. Furthermore, 5G4 was the only antibody tested that labelled astrocytic αSyn [14]. Altay and colleagues similarly reported that 5G4 and other αSyn antibodies targeting the central NAC region of the αSyn, but not SYNO4 detected astrocytic αSyn [15]. Astrocytic αSyn accumulation is still a relatively new aspect of LBD neuropathology but increasingly recognised in large-scale studies [16]. Here, we demonstrate that astrocytic αSyn is detectable in paraffin-embedded tissue stored for up to 78 years, in line with previous studies of recently deceased patients [15, 17] and in cases with prolonged fixation times up to 14 years [14]. Our study also further confirms that Lewy pathology is often accompanied by astrocytic αSyn.

Due to the broad range of the groups and fixation times, we only tested one antibody per protein aggregate. Additional studies are needed to evaluate the efficacy of other antibodies targeting αSyn, Aβ, and hyperphosphorylated tau. Furthermore, although protocols were optimised as part of this study, our aim was not to present specific optimisations, but to demonstrate the staining quality and immunoreactivity achievable with optimised protocols. Antigen retrieval with FA and heat has proved to be essential for unmasking the antigens, particularly for αSyn and Aβ [8, 10, 11, 17, 18]. As part of the protocol optimisation process, antigenicity was remarkably reduced when staining for αSyn without FA treatment (data not shown). Based on previous studies, 5G4 was chosen for detection of αSyn, and our results confirm its utility even after fixation times up to 78 years. Similarly, the 4G8 antibody proved robust for detecting senile plaques across different antigen retrieval methods in archival brain tissue with prolonged fixation times, consistent with previous reports [10].

For tau, the AT8 antibody showed variable effects depending on the type of aggregate. Intracellular inclusions in long-fixed tissue maintained similar staining intensity and quality as short-fixed tissue, whereas NTs showed reduced labelling, consistent with previous observation by Pikkarainen and colleagues who also identified limited change in the number of NFTs [9, 11]. Earlier work by Dwork and colleagues reported good labelling of the Alz 50 epitope after 10 years of fixation, but reduced immunoreactivity with longer fixation times, and contrary to our findings, the immunosignal was completely absent after 30 years of fixation [19]. This discrepancy highlights that both the choice of antibody and protocol optimisation strongly impact the labelling of protein aggregates after prolonged fixation times [11].

This study has several limitations. First, sampling of new blocks depended on the original sampling at the time of neuropathological evaluations (1946-1980), and for some cases, sampling could not be performed completely adjacent to the original site. Thus, some biological variation could have impacted our results, as decreased pathology in new blocks could reflect regional differences rather than antigen loss. Second, the burden of pathology for each case was not known prior to inclusion into the current study; some groups were positive for all three pathologies, whereas others were a mix of negative and positive cases of the types of pathology investigated. The 1970s group showed higher semi-quantitative scores for LBs and LNs, likely reflecting group composition rather than a preservation effect. Sample sizes, particularly for the oldest groups regarding αSyn, were limited. It was challenging to add more αSyn-positive cases fitting into the criteria of the decades as approximately half the cases we had identified as potential αSyn-positive cases were negative when screened for αSyn in the SN.

Across the different pathologies and their corresponding antigens, a slight decrease was observed in the new blocks compared to the original blocks. However, the differences between the preservation categories were smallest in the 1940s and the 1970s groups. This suggest that the reduction in antigenicity of these protein aggregates may reach a plateau after a few years of fixation, and that additional years or even decades of fixation do not further reduce the antigenicity proportional to the duration of fixation. Visualisation of antigens is dependent on both the antibody and the pretreatment method, as reported by other studies [8, 10, 11, 14, 15, 17, 20]. Thus, investigating antigen decay in very long-fixed tissue is challenging, since negative IHC staining result may be due to a suboptimal antibody or antigen retrieval protocol rather than true antigen loss. Due to the limited information available on fixative type, postmortem delay, and fixation times of original blocks, we cannot comment on the influence of these factors individually. However, as we performed same-case comparisons, any influence from these variables most likely had relatively minor impact on the present findings.

Examples of “false-negative” and “false-positive” IHC staining results were noticed when comparing the immunoreactivity of new blocks to original blocks (see coloured sections of **Table 4**). A few cases showed discrepant results, which could have several possible explanations. One is a true pathology-negative finding in one block due to unknown technical differences. The use of an automated immunostainer in the present study should reduce variability in the IHC procedure by ensuring uniform application of each protocol step and improving reproducibility, therefore the explanation is likely to lie elsewhere. Another possibility is slight anatomical differences between sampled regions, such that negative staining in one block and positive staining in another may reflect the true distribution of pathology. For most cases showing discrepant IHC results, this occurred for only one antibody in one region per case. In these cases, the remaining regions as well as the immunoreactivity of the other two antibodies showed adequate-to-good IHC results. Postmortem delay is one factor that can affect antigenicity. One case presented discrepant results across all regions assessed for tau pathology, whereas αSyn pathology was well-labelled in both original and new blocks in all regions. However, the postmortem delay in this formalin-fixed case was 25 hours, which is within commonly accepted limits. Taken together with the semi-quantitative analysis, this could indicate that the tau antigenicity is more vulnerable for prolonged fixation and possibly other factors, making epitope unmasking and antigen retrieval more difficult. The effects of other pre-sectioning factors, apart from prolonged fixation, lie beyond the scope of this study, as the necessary information to explore these parameters were unavailable to us.

Overall, our results indicate that when studying protein aggregates of AD and LBD, it is possible to obtain adequate-to-good staining in most cases irrespective of postmortem delay ranging from 6 to 113 hours, storage times of up to 78 years, and fixation times of up to 78 years. Future research could include testing a panel of antibodies against αSyn, tau, and Aβ to investigate limitations in the range of available antibodies when studying archival brain tissue. In addition, IHC labelling of neuroinflammatory markers, as well as astrocytic αSyn using antibodies against various truncated forms of αSyn could be explored in tissue with very prolonged fixation times. Furthermore, tissue clearing, spatial transcriptomics, and the feasibility of DNA and RNA analyses in both types of archival tissue could also be investigated.

Here, we have shown that, with appropriate antigen retrieval methods and antibody selection, archival brain tissue stored either as paraffin-embedded blocks or in fixative for up to 78 years can be successfully used to evaluate the defining neuropathological features of LBD and AD.

## Data Availability

All data produced in the study are available upon reasonable request to the authors.

## Abbreviations

Aβ: amyloid-beta
AD: Alzheimer’s Disease
αSyn: alpha-synuclein
CAA: cerebral amyloid angiopathy
Ent Cx: entorhinal cortex
FA: formic acid
FFPE: formalin-fixed paraffin-embedded
IHC: immunohistochemical
LB: Lewy body
LBD: Lewy Body Disease
LN: Lewy neurite
NFT: neurofibrillary tangle
NT: neuropil thread
PAG: periaqueductal grey
PD: Parkinson’s Disease
ROI: region of interest
SN: substantia nigra
SP: senile plaque
TO Cx: temporo-occipital cortex

## Acknowledgements

We would like to acknowledge the donors of the Danish Brain Collection and BRIDGE at Southern University of Denmark for running the brain bank thereby making this study possible.

